# Attitudes and Perceptions of Generative Artificial Intelligence Chatbots in the Peer Review of Traditional, Complementary, and Integrative Medicine Research: A Large-Scale, International Cross-Sectional Survey

**DOI:** 10.64898/2026.11.12.26366211

**Authors:** Jeremy Y. Ng, Jamie Tan, Karthik Adapa, Prashant Kumar Gupta, Madhur Gupta, Shao Li, Darshan H. Mehta, Melinda Ring, Manisha Shridhar, João Paulo Souza, Tetsuhiro Yoshino, Myeong Soo Lee, Holger Cramer

**Author notes:** Corresponding Author Details: Name: Dr. Jeremy Y. Ng, MSc, PhD, Address: Institute of General Practice and Interprofessional Care, University Hospital Tübingen, Osianderstr. 5, 72076 Tübingen, or.

## Abstract

**Background:** Generative artificial intelligence (GenAI) chatbots are being explored for their potential to support the peer review process by improving efficiency and analytical capacity. While early applications show promise, concerns regarding ethics, privacy, and research integrity remain, and their use within traditional, complementary, and integrative medicine (TCIM) remains underexplored. This study examines TCIM researchers’ attitudes and perceptions regarding the use of GenAI chatbots in peer review through a large-scale international survey.

**Methods:** Corresponding authors of TCIM-related publications indexed in Ovid MEDLINE were invited to participate in an online, anonymous, international cross-sectional survey distributed via SurveyMonkey. The questionnaire collected data on demographic characteristics, the perceived importance of GenAI in peer review, the likelihood of adoption, and the perceived benefits and concerns.

**Results:** A total of 889 eligible respondents participated. Overall, 40.25% (n=285/708) reported a positive perception of GenAI in peer review, and 41.67% (n=295/708) considered it important to the field. 34.24% researchers reported being likely (n=253/739) or 18.27% (135/739) very likely (totaling 52.51%) to use GenAI in peer review, highlighting a gap between perceived importance and intended adoption. Identified benefits included improved efficiency (50.72%, n=350/690), reduced reviewer workload (51.16%, n=353/690), and generation of constructive feedback (46.30%, n=319/689), reflecting respondents selecting “agree.” Concerns included bias (49.85%, n=333/668), lack of transparency (42.86%, n=285/665), inaccuracies (49.40%, n=329/666), accountability challenges (45.20%, n=301/666), and data privacy risks (36.05%, n=239/663). Most respondents believed that peer reviewers require either some training (43.91%, n=310/706) or a lot of training (40.93%, n=289/706) to use GenAI chatbots effectively in peer review.

**Conclusion:** Respondents reported perceived benefits of GenAI in peer review, alongside notable concerns regarding its use. While opinions were mixed, findings suggest both perceived utility and apprehension about potential risks. Further work is needed to understand these perspectives better and to explore how GenAI can be responsibly integrated into peer-review processes.

## Background

### Introduction to Generative Artificial Intelligence in Peer Review

Artificial intelligence (AI) technologies are becoming increasingly embedded within academic research and editorial processes, including peer review, offering tools that can enhance the efficiency, consistency, and objectivity of various tasks [1]. Conventional AI systems, including machine learning models, have already supported activities such as plagiarism detection, image verification, and statistical integrity checks [2]. More recently, GenAI chatbots, a category of AI that includes large language models such as OpenAI’s ChatGPT and Google Gemini, show particular potential for assisting peer reviewers by summarizing manuscripts, evaluating coherence, and suggesting improvements to writing quality [3]. These technologies may streamline peer-review workflows, foster consistency in critiques, and expand access to advanced analytical support, especially for reviewers in low-resource settings [1]. They may also help reviewers communicate their assessments more clearly, which may be particularly valuable for non-native-English-speaking reviewers [1].

However, concerns have emerged regarding the integration of GenAI chatbots into the peer review process. These include the possibility of factual errors, opacity in how outputs are generated, amplification of existing biases, issues related to manuscript confidentiality, data security, intellectual property, and ambiguity in responsibility and accountability for GenAI chatbot-assisted reviews [1]. Overdependence on such tools may also impair reviewers’ critical thinking and analytical reasoning, which is particularly concerning for early-career or less experienced reviewers [1, 4]. It is essential to recognize both the benefits and risks of GenAI chatbots to ensure their responsible and equitable use in the peer review of scientific research.

### The Role of Generative Artificial Intelligence in Supporting Peer Review of Traditional, Complementary, and Integrative Medicine Research

Traditional, complementary, and integrative medicine (TCIM) comprises a wide range of practices, including Ayurvedic medicine, acupuncture, herbal medicine, and mind-body therapies, many of which operate outside conventional biomedical paradigms [5]. Traditional medicine encompasses culturally rooted knowledge and practices, beliefs, and skills used to promote health and prevent, diagnose, and treat physical and mental illnesses [6]. The World Health Organization (WHO) defines complementary medicine as “additional healthcare practices that are not part of a country’s mainstream medicine” [7]. Comparatively, integrative medicine “is an interdisciplinary and evidence-based approach to health and well-being by using a combination of biomedical and traditional and/or complementary medical knowledge, skills, and practices” [7].

Conducting rigorous research (and therefore peer review) in the TCIM field poses particular challenges due to the diversity of therapeutic systems, variation in research designs, and the methodological complexity of evaluating non-conventional interventions [8–10]. Researchers have begun using AI technologies in TCIM research [11–13]. GenAI chatbots may also offer valuable support to peer reviewers of TCIM research by helping to synthesize detailed content, assess methodological soundness, and enhance the clarity and structure of peer review reports, while simultaneously reducing the burden on reviewers and benefitting journals that face challenges in recruiting reviewers with relevant expertise [1, 14]. These GenAI chatbots also have the potential to address global inequities in the peer-review process by providing language support and analytical assistance to reviewers from diverse backgrounds and regions [15]. As the volume of TCIM literature continues to grow and as more patients incorporate TCIM into their care, leveraging AI technologies may become increasingly important for improving the quality, consistency, and accessibility of peer review across the TCIM research landscape [1, 16].

### Gap in Knowledge

Although GenAI tools hold considerable promise for transforming peer review, little is currently known about how peer reviewers of TCIM research perceive their use in this context. While it has been established that peer reviewers of the medical literature (in general) are using GenAI chatbots to assist them with peer review [17], there is a lack of evidence on peer reviewers’ attitudes toward GenAI chatbots, including perceived advantages, limitations, and ethical implications, specifically in the context of TCIM research. Understanding these views is essential to inform the thoughtful development and integration of GenAI chatbots in ways that respect the distinctive values, practices, and challenges of the TCIM community. Addressing key issues such as transparency, fairness, and reviewer accountability is also necessary to build confidence and promote responsible use of these technologies among peer reviewers [16].

### Relevance and Impact

Examining the perspectives of peer reviewers of TCIM research on the use of GenAI chatbots in the peer review process is both timely and important. These tools may offer benefits such as streamlining review tasks, providing analytical support, and improving the quality and consistency of reviewer feedback [16, 18, 19]. GenAI chatbots could also help address some of the complexities of reviewing TCIM manuscripts, including interdisciplinary content, unfamiliar methodologies, and varied terminology, by assisting with content synthesis, translation, and interpretation [5, 20–22]. However, ethical issues, including data security, potential bias in algorithmic output, and questions of reviewer responsibility, must be carefully considered to ensure appropriate implementation [16]. Intellectual property concerns, particularly those involving traditional knowledge embedded in TCIM research, further highlight the need for clear ethical safeguards to prevent misuse or misappropriation by AI systems [16, 23]. The findings of this study will help shape the appropriate use of GenAI tools in TCIM peer review and support efforts to improve the quality and transparency of scholarly communication in the field. In summary, this study seeks to address the current knowledge gap by investigating peer reviewers’ attitudes and perceptions toward the use of GenAI chatbots in reviewing TCIM research. The insights gained will inform best practices for the responsible adoption of GenAI in peer review and contribute to broader efforts to advance quality in academic publishing.

## Methods

### Open Science Statement

A cross-sectional, web-based survey was conducted among TCIM researchers identified as corresponding authors of subject-specific articles indexed in MEDLINE. The study protocol was registered on the Open Science Framework (OSF) [24]. Before initiating any data collection, ethics approval was sought from the University Hospital Tubingen Research Ethics Board (REB #: 543/2025BO2). All study-related materials and data sets were uploaded to OSF and can be accessed at https://doi.org/10.17605/OSF.IO/M6SGP. Upon completion of data collection and analysis, the findings were compiled into a manuscript and shared as a preprint before submission to a peer-reviewed journal. The final manuscript was developed in accordance with the Strengthening the Reporting of Observational Studies in Epidemiology (STROBE) guidelines for cross-sectional studies [25] and the Checklist for Reporting Results of Internet E-Surveys (CHERRIES) [26].

### Sampling Framework

We searched Ovid MEDLINE (ALL, 1946 to July 16, 2025) to identify relevant literature on TCIM published between June 1, 2015, and June 1, 2025. The strategy combined controlled vocabulary (MeSH) terms and keywords to capture a broad range of TCIM domains, including acupressure, acupuncture, chiropractic, complementary and alternative therapies, Chinese herbal medicine, electroacupuncture, herbal medicine, integrative medicine, applied kinesiology, osteopathic medicine and manipulation, Ayurvedic medicine, traditional Chinese and East Asian medicine, mind–body therapies, naturopathy, phytotherapy, medicinal plants, and yoga. To improve specificity, we excluded publication types that do not represent original research, such as commentaries, editorials, letters, news items, biographies, conference proceedings, case reports, and retractions. Finally, results were restricted to English-language publications within the specified date range. Based on this search strategy, it can be assumed that a significant proportion of the corresponding authors of these articles are also peer reviewers of TCIM research. PubMed Identifier (PMID) numbers from the resulting articles were exported from MEDLINE as.csv files and processed using an R script (based on the 2019 easyPubMed and RedEye packages) to extract author names, affiliated institutions, and email addresses [27, 28]. Once the PMIDs were imported into R, the corresponding author email addresses were extracted, and the data were subsequently cleaned for errors or duplicates. Additionally, extracted email addresses were manually checked for truncation and verified with a hex recorder developed in PyCharm before survey distribution [28]. Authors who met the inclusion criteria as outlined in the sampling framework were invited to participate in an anonymous online survey [24].

### Participant Recruitment

Only TCIM researchers identified through the sampling framework were invited to take part in the study. These potential participants were contacted by email and invited to complete the survey voluntarily and anonymously. Emails were distributed via SurveyMonkey and included information outlining the study’s objectives, along with a link to the informed consent form [29]. Participation was fully voluntary, and no compensation was offered. Participants were provided with an informed consent form before accessing the survey, and consent to participate was implied by completion of the survey (https://osf.io/7vqj4/files/wj5k2). Individuals with multiple publications were only invited to complete the survey once, based on the most recent article identified in the search strategy. The first email invitation was sent on March 17, 2026, and three weekly reminders were sent to participants on March 24, March 31, and April 7, 2026. The survey remained open after the last reminder and was closed on April 25, 2026. Duplicate entries based on identical name and email combinations were removed before analysis. Copies of the initial and follow-up email invitations are available online at https://osf.io/7vqj4/files/vsk29. Respondents were able to skip any survey questions they preferred not to answer. No personal information was collected, and participants’ IP addresses were not retained. Informed consent covering the collection, storage, use, and sharing of study data was obtained from all participants before the survey, and participants could withdraw at any time.

### Survey Design

The survey was constructed on the SurveyMonkey platform [29]. Participants received an email containing a link to the informed consent form and survey. Consent to participate was implied by completion of the survey. The questionnaire consisted of 31 questions across 13 pages, beginning with multiple-choice questions and followed by general demographic items. These demographic questions were adapted from prior studies in the TCIM field [30, 31]. After completing the demographic section, participants responded to items assessing their familiarity with, attitudes toward, and experience using GenAI chatbots in the context of peer review. These questions addressed different aspects of the peer review process, including evaluating methods, interpreting findings, assessing clarity and structure, and providing constructive feedback. The GenAI chatbot-related items were based on prior research examining attitudes and perceptions toward AI across various scholarly contexts [30, 31]. The full survey instrument is provided in the published protocol [24].

### Data Analysis

This study did not involve the testing of formal hypotheses. Responses were included if the participant provided consent and met the eligibility criteria. Therefore, eligible partial survey responses were retained and included in the final analysis. Quantitative data were analyzed using descriptive statistics, including frequencies and percentages, generated through SurveyMonkey [29]. To descriptively compare responses across participant groups, responses were stratified by sex, age, geographic location, career stage, and whether TCIM was the respondent’s primary or secondary research focus. Additionally, responses were stratified by age, sex, location of employment, career stage, and whether TCIM represented a primary or secondary research focus using SurveyMonkey [29]. Where sufficient qualitative data were available, responses were analyzed using an inductive thematic content analysis. Two members of the research team independently coded the qualitative responses in duplicate following a pilot coding exercise. The coding approach relied exclusively on inductive methods, allowing themes to emerge directly from the raw data without imposing pre-existing theoretical frameworks. They engaged in iterative discussions to refine the coding structure and achieve consensus, ensuring analytic rigour. The process emphasized generating mutually exclusive, clearly defined categories, guided by constant comparison and category refinement to ensure analytic rigour and trustworthiness [32, 33].

## Results

Survey invitations were distributed across multiple recruitment waves, resulting in a total of 63,666 invitations sent to corresponding authors. Of the survey invitations distributed, 24,087 (37.83%, n=24,087/63,666) were opened, 32,066 (50.37%, n=32,066/63,666) were not opened, 6,948 (10.91%, n=6,948/63,666) bounced, and 565 recipients (0.89%, n=565/63,666) opted out. In total, 2,084 recipients (3.27%, n=2,084/63,666) clicked through the survey link. Of the 2,084 participants who accessed the survey, 1,088 (1.71%, n=1,088/63,666) submitted the first survey page or consented to participate, corresponding to a participation rate of 52.21% (1,088/2,084). Of the 1,088 survey responses received, 199 were excluded as ineligible, leaving 889 eligible participants. Among these eligible respondents, 875 completed the survey, resulting in a completion rate of 98.43%. This indicates a CHERRIES completion rate of 80.42% (875/1,088). Respondents were permitted to skip individual survey items; therefore, the number of responses varied across questions. Incomplete survey questions were retained for analysis. Raw data of the survey is available online at https://osf.io/7vqj4/files/du9ma. On average, the survey took 9 minutes and 37 seconds to complete.

Demographic data can be found in **Table 1**. Most respondents were male (53.82%, n=465/864), while 44.21% identified as female (n=382/864). The largest age group was 45–54 years (30.29%, n=262/865), followed closely by 35–44 years (29.94%, n=259/865), and 55–64 years (19.19%, n=166/865). Most respondents did not identify as being part of a visible minority group, defined as a non-dominant ethnic group in their country of employment (78.79%, n=680/863). Respondents primarily worked in South-East Asia (29.65%, n=255/860), Europe (27.67%, n=238/860), and the Americas (21.63%, n=186/860). Given the option to select more than one role, most respondents identified as faculty members at a university or academic institution (71.62%, n=540/754), followed by academic research staff (22.02%, n=166/754) and clinician researchers (16.98%, n=128/754). The largest proportion of respondents were senior researchers (69.51%, n=522/751). Respondents reported publishing a wide range of peer-reviewed TCIM research articles, from 0 to over 500, with most reporting fewer than 100. Some respondents reported a range rather than a specific number of published articles. These responses were excluded from the calculations presented in **Table 1**. Results stratified by age, sex, location of employment, career stage, and whether TCIM represented a primary or secondary research interest are available at https://osf.io/7vqj4/files/6zkmj.

**Table 1:** TCIM Peer Reviewers’ Demographics.

| **Sex** | **Responses** | **Percentage** |
|---|---|---|
| Female | 382 | 44.21% |
| Male | 465 | 53.82% |
| Prefer not to say | 14 | 1.62% |
| Prefer to self-describe | 3 | 0.35% |
| Total | 864 | 100.00% |
| **Age** | **Responses** | **Percentage** |
| Under 18 | 0 | 0.00% |
| 18-24 | 1 | 0.12% |
| 25-34 | 74 | 8.55% |
| 35-44 | 259 | 29.94% |
| 45-54 | 262 | 30.29% |
| 55-64 | 166 | 19.19% |
| 65 or older | 94 | 10.87% |
| Prefer not to say | 9 | 1.04% |
| TOTAL | 865 | 100.00% |
| **Minority Group Status** | **Responses** | **Percentage** |
| Yes | 106 | 12.28% |
| No | 680 | 78.79% |
| Prefer not to say | 77 | 8.92% |
| TOTAL | 863 | 100.00% |
| **Country of Employment** | **Responses** | **Percentage** |
| Africa | 74 | 8.60% |
| Americas | 186 | 21.63% |
| Eastern Mediterranean | 38 | 4.42% |
| Europe | 238 | 27.67% |
| South-East Asia | 255 | 29.65% |
| Western Pacific | 69 | 8.02% |
| TOTAL | 860 | 100.00% |
| **Current Position (Select all that apply)** | **Responses** | **Percentage** |
| Faculty member at a university/academic institution | 540 | 71.62% |
| Academic research staff (e.g., research coordinator at a university/academic institution) | 166 | 22.02% |
| Government researcher | 26 | 3.45% |
| Clinician researcher | 128 | 16.98% |
| Pharmaceutical industry/company employee | 4 | 0.53% |
| Scholarly communications employee (e.g., scholarly journals/publishing, medical writing) | 13 | 1.72% |
| Consultant (e.g., own or employed by research consulting firm) | 25 | 3.32% |
| Third sector (e.g., NGO, non-profit) | 13 | 1.72% |
| Other (please specify) | 34 | 4.51% |
| TOTAL | 754 | 100.00% |
| **Career Stage** | **Responses** | **Percentage** |
| Early career researcher (<5 years ) | 74 | 9.85% |
| Mid-career researcher (5-10 years) | 155 | 20.64% |
| Senior researcher (>10 years) | 522 | 69.51% |
| TOTAL | 751 | 100.00% |
| **Primary Research Area** | **Responses** | **Percentage** |
| Clinical research | 325 | 45.52% |
| Preclinical research – in vivo | 105 | 14.71% |
| Preclinical research – in vitro | 96 | 13.45% |
| Health systems research | 41 | 5.74% |
| Health services research | 55 | 7.70% |
| Methods research | 66 | 9.24% |
| Epidemiological research | 26 | 3.64% |
| Other (please specify) | 65 | 9.10% |
| TOTAL | 714 | 100.00% |
| **Number of Published Research Articles** | **Responses** | **Percentage** |
| Under 100 | 635 | 90.20% |
| 100-199 | 44 | 6.25% |
| 200-299 | 11 | 1.56% |
| 300-399 | 9 | 1.28% |
| 400 or more | 5 | 0.71% |
| TOTAL | 704 | 100% |
| TCIM Research Focus | Responses | Percentage |
| Primary | 374 | 50.34% |
| Secondary | 369 | 49.66% |
| TOTAL | 743 | 100.00% |
| **Most TCIM Research Conducted in:** | **Responses** | **Percentage** |
| Mind-body therapies (e.g., meditation, biofeedback, hypnosis, yoga, tai chi, imagery, creative outlets) | 130 | 17.66% |
| Biologically based practices (e.g., vitamins and dietary supplements, botanicals, special foods, and diets) | 249 | 33.83% |
| Manipulative and body-based practices (massage, chiropractic therapy, reflexology) | 73 | 9.92% |
| Biofield therapy (e.g., Reiki, therapeutic touch) | 11 | 1.49% |
| Whole medical systems (e.g., Ayurvedic medicine, East Asian medicine, acupuncture, homeopathy, naturopathic medicine) | 157 | 21.33% |
| Other, please specify (e.g., methodology, TCIM professions, etc.) | 116 | 15.76% |
| TOTAL | 736 | 100.00% |

Respondents’ familiarity with GenAI chatbots and views on their use in the peer review process are shown in **Table 2**. Most respondents reported being “familiar” (61.52%, n=454/738) or “very familiar” (18.83%, n=139/738) with GenAI chatbots. In a multiple-response question (select all that apply), OpenAI’s ChatGPT was the most frequently reported GenAI chatbot used for any purpose (85.28%, n=614/720 respondents), followed by Google Gemini (formerly Google Bard) (50.69%, n=365/720 respondents) and Microsoft Copilot (formerly Bing Chat) (34.86%, n=251/720 respondents). OpenAI’s ChatGPT was also the most commonly used GenAI chatbot for peer review-related purposes (37.78%, n=269/712 respondents), while more than half of respondents (56.46%, n=402/712) reported never using a GenAI chatbot for peer review-related purposes. Regarding future use, 34.24% (n=253/739) of respondents reported being “likely” and 18.27% (n=135/739) “very likely” to use GenAI chatbots for peer review in the future. Similarly, most respondents felt that GenAI chatbots would be “important” (41.67%, n=295/708) or “very important” (37.99%, n=269/708) in the future of peer review. Overall, respondents generally viewed the impact of GenAI chatbots on the future of peer review positively, with 40.25% (n=285/708) reporting positive perceptions and 21.47% (n=152/708) reporting very positive perceptions. In contrast, 15.82% (n=112/708) perceived the impact negatively and 5.79% (n=41/708) very negatively.

**Table 2:** TCIM Peer Reviewers’ Familiarity, Use, Training, and Perceptions of GenAI in the Peer Review Process.

| **How familiar are you with GenAI chatbots?** | **Responses** | **Percentage** |
|---|---|---|
| Very familiar | 139 | 18.83% |
| Familiar | 454 | 61.52% |
| Unfamiliar | 96 | 13.01% |
| Very unfamiliar | 31 | 4.20% |
| I am not sure | 18 | 2.44% |
| Total | 738 | 100.00% |
| **Which GenAI chatbots have you used in the past for any purpose? (Select all that apply)** | **Responses** | **Percentage** |
| ChatGPT | 614 | 85.28% |
| Google Gemini (formerly Google Bard) | 365 | 50.69% |
| Meta AI | 84 | 11.67% |
| Microsoft Copilot (formerly Bing Chat) | 251 | 34.86% |
| I have never used a GenAI chatbot | 60 | 8.33% |
| Other (please specify) | 115 | 15.97% |
| Total | 720 | 100.00% |
| **Which GenAI chatbots have you used in the past for peer review? (Select all that apply)** | **Responses** | **Percentage** |
| ChatGPT | 269 | 37.78% |
| Google Gemini (formerly Google Bard) | 101 | 14.19% |
| Meta AI | 20 | 2.81% |
| Microsoft Copilot (formerly Bing Chat) | 37 | 5.20% |
| I have never used a GenAI chatbot for purposes relating to the peer review process | 402 | 56.46% |
| Other (please specify) | 42 | 5.89% |
| TOTAL | 712 | 100.00% |
| **How likely are you to use GenAI chatbots for peer review in the future?** | **Responses** | **Percentage** |
| Very likely | 135 | 18.27% |
| Likely | 253 | 34.24% |
| Unlikely | 108 | 14.61% |
| Very unlikely | 129 | 17.46% |
| I'm not sure | 114 | 15.43% |
| Total | 739 | 100.00% |
| **Has your research institution provided any education/training on appropriate GenAI use in peer review?** | **Responses** | **Percentage** |
| Yes, and I have taken it | 92 | 12.48% |
| Yes, but I have not taken it | 92 | 12.48% |
| No | 431 | 58.48% |
| I am not sure | 122 | 16.55% |
| Total | 737 | 100.00% |
| **Has your research institution implemented any policies surrounding the use of GenAI chatbots in the peer review process?** | **Responses** | **Percentage** |
| Yes | 88 | 12.05% |
| No | 401 | 54.93% |
| I’m not sure | 241 | 33.01% |
| TOTAL | 730 | 100.00% |
| **How much training and education do you think researchers need in order to effectively use GenAI chatbots in the peer review process?** | **Responses** | **Percentage** |
| A lot | 289 | 40.93% |
| Some | 310 | 43.91% |
| Very little | 45 | 6.37% |
| None | 17 | 2.41% |
| I am not sure | 45 | 6.37% |
| TOTAL | 706 | 100.00% |
| **Would you be interested in learning more/receiving training about how to use GenAI chatbots in the peer review process?** | **Responses** | **Percentage** |
| Yes | 460 | 65.43% |
| No | 77 | 10.95% |
| Maybe | 166 | 23.61% |
| TOTAL | 703 | 100.00% |
| **How important do you think GenAI chatbots will be in the future of peer review?** | **Responses** | **Percentage** |
| Very important | 269 | 37.99% |
| Important | 295 | 41.67% |
| Unimportant | 36 | 5.08% |
| Very unimportant | 13 | 1.84% |
| I am not sure | 95 | 13.42% |
| TOTAL | 708 | 100.00% |
| **In general, how do you perceive the potential impact of GenAI chatbots on the future of peer review?** | **Responses** | **Percentage** |
| Very positively | 152 | 21.47% |
| Positively | 285 | 40.25% |
| Negatively | 112 | 15.82% |
| Very negatively | 41 | 5.79% |
| I am not sure | 118 | 16.67% |
| TOTAL | **708** | 100.00% |

A large proportion of respondents reported that their research institution did not provide education or training on the appropriate use of GenAI chatbots in peer review (58.48%, n=431/737). Smaller proportions indicated that their institution offered training, but they had not taken it (12.48%, n=92/737), or that they had completed institutional training (12.48%, n=92/737). Additionally, most respondents reported that their institution had not implemented policies regarding the use of GenAI chatbots in the peer-review process (54.93%, n=401/730). In comparison, only 12.05% (n=88/730) indicated that such policies were in place.

The largest proportion of respondents believed that peer reviewers require some training and education to effectively use GenAI chatbots in peer review (43.91%, n=310/706). In comparison, 40.93% (n=289/706) believed that a lot of training is required. Furthermore, most respondents expressed interest in receiving additional training on using GenAI chatbots in the peer review process (65.43%, n=460/703), while 23.61% (n=166/703) responded “maybe”.

Peer reviewers were also asked to rate their personal use of GenAI chatbots at different stages of the peer review process on a five-point scale from never to always (see **Figure 1**). Overall, most respondents reported never using GenAI chatbots for most peer-review-related tasks. The highest proportions of respondents reporting “never” using GenAI chatbots were for identifying potential conflicts of interest (73.61%, n=516/701), identifying fabrication or falsification of data (69.89%, n=492/704), and assisting with assessing ethical considerations or compliance issues (69.33%, n=486/701). Similarly, most respondents reported never using GenAI chatbots to identify plagiarism or duplication in manuscripts (62.98%, n=444/705), to interpret statistical analyses and critique their validity (61.11%, n=429/702), and to evaluate the suitability of research methodology (59.60%, n=419/703).

**Figure 1.**
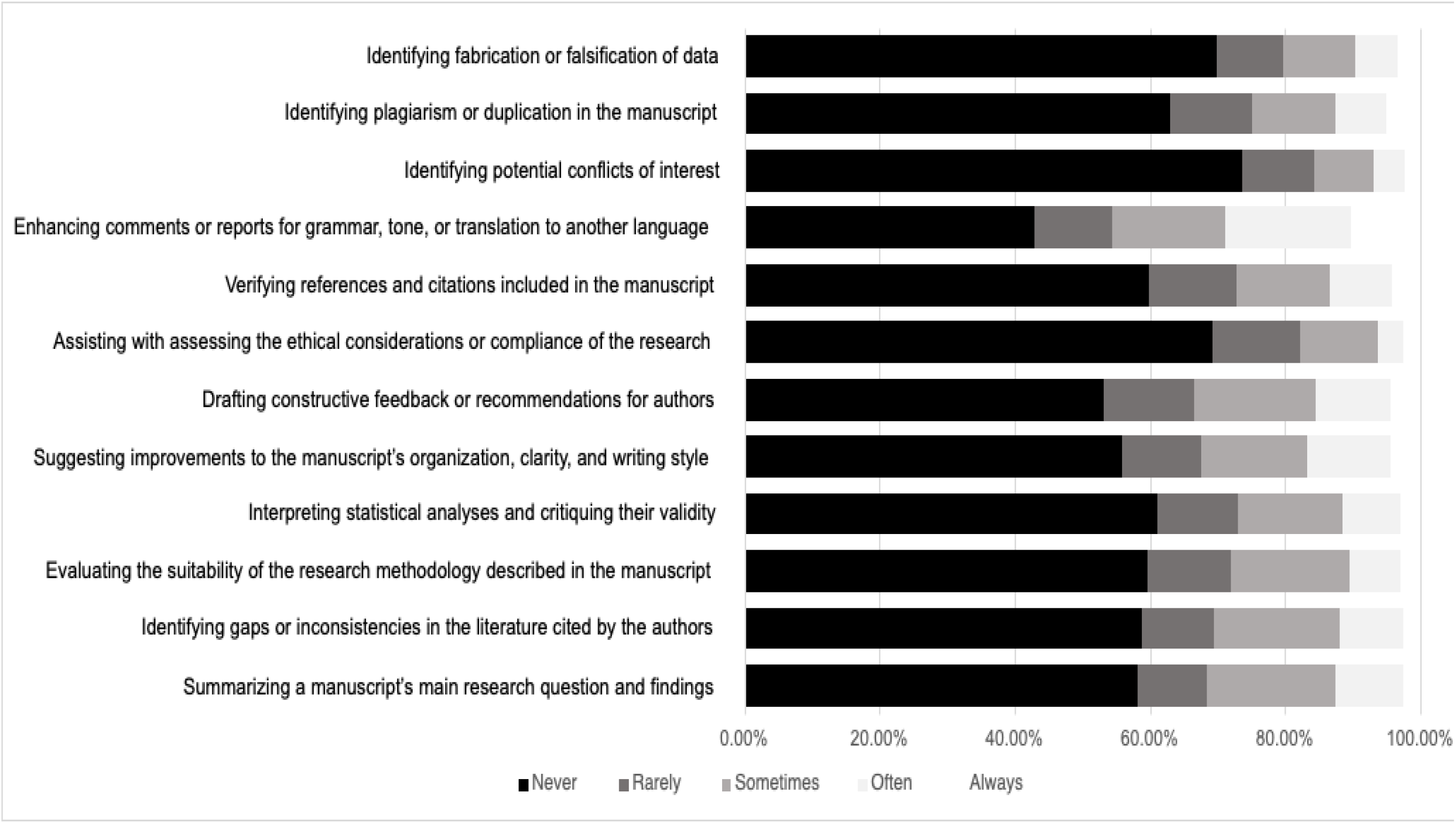
TCIM Peer Reviewers’ Self-Reported Use of GenAI Chatbots Across Different Stages of the Peer Review Process.

In contrast, respondents reported comparatively greater use of GenAI chatbots for language-related and organizational tasks. Enhancing comments or reports for grammar, tone, or translation had the highest proportion of respondents reporting “often” using GenAI chatbots (18.58%, n=131/705), while 10.21% (n=72/705) reported “always” using them for this purpose. Respondents also reported relatively higher use of GenAI chatbots for summarizing a manuscript’s main research question and findings, with 19.03% (n=134/704) reporting “sometimes,” 9.94% (n=70/704) reporting “often,” and 2.56% (n=18/704) reporting “always.” Similar trends were observed in identifying gaps or inconsistencies in the cited literature and drafting constructive feedback or recommendations for authors, with “sometimes” the most frequently selected response among respondents who reported using GenAI chatbots for these tasks. Overall, the findings suggest that respondents were more accepting of using GenAI chatbots for supportive writing and editing tasks than for evaluative or integrity-related aspects of peer review.

Using a five-point scale ranging from strongly disagree to strongly agree, respondents rated their perceptions of the proposed benefits of using a GenAI chatbot in the peer review process (see **Figure 2**). Respondents most frequently agreed that GenAI chatbots could reduce the workload of peer reviewers (51.16%, n=353/690), increase the speed and efficiency of conducting reviews (50.72%, n=350/690), and improve the quality and readability of peer review reports (48.33%, n=333/689). Respondents also agreed that GenAI chatbots could help identify errors or inconsistencies in manuscripts (40.43%, n=279/690) and support interdisciplinary collaboration by helping reviewers better understand unfamiliar research areas (40.61%, n=279/687).

**Figure 2.**
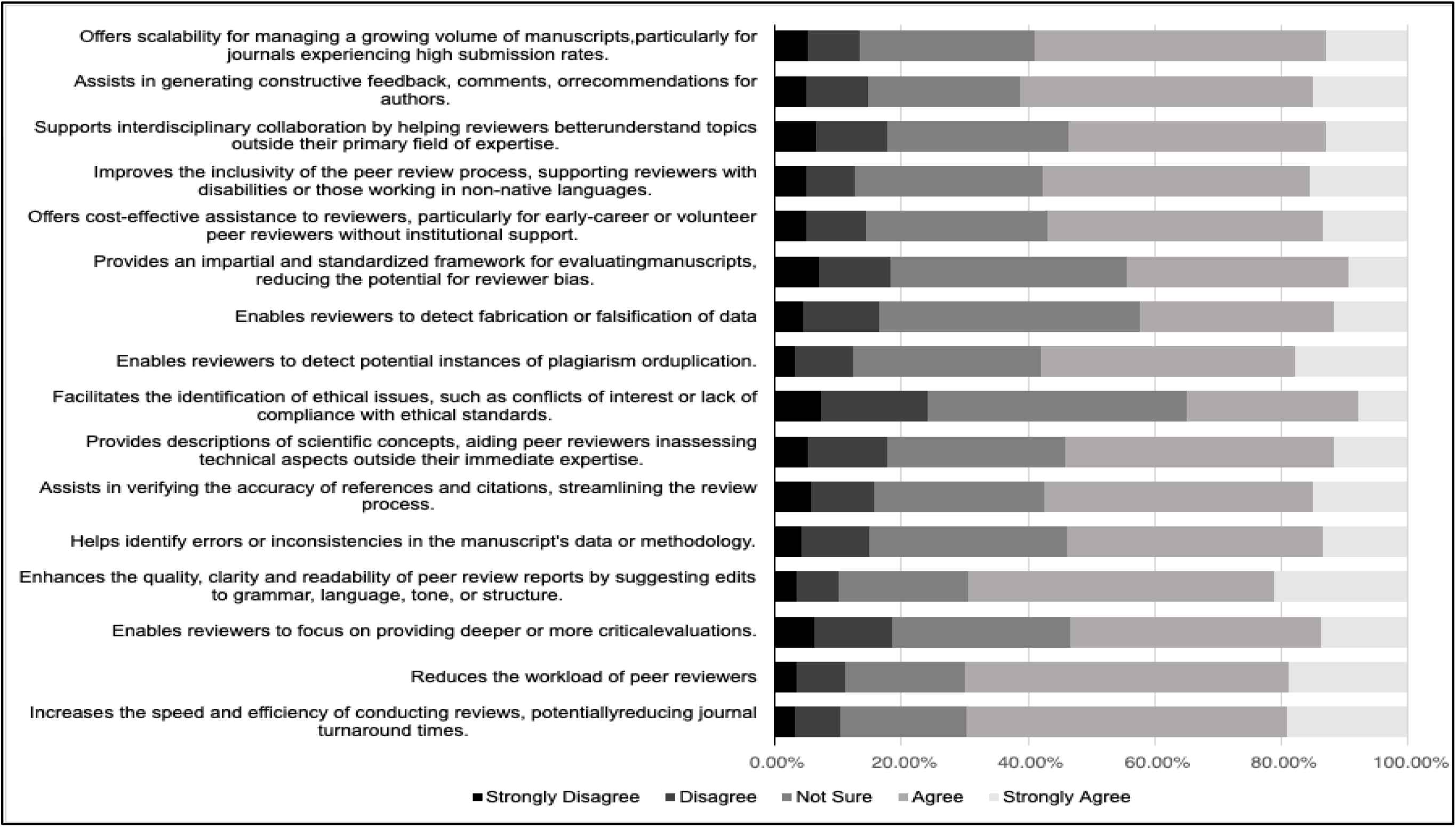
TCIM Peer Reviewers’ Perceptions of the Proposed Benefits of GenAI Chatbot Use in the Peer Review Process.

However, respondents also expressed substantial concerns about the use of GenAI chatbots in peer review (**Figure 3**). Respondents most frequently agreed that GenAI chatbots may introduce bias (49.85%, n=333/668), struggle to interpret complex scientific reasoning and nuanced arguments (47.67%, n=317/665), and raise ethical and legal concerns related to transparency, privacy, and consent (41.38%, n=276/667). Concerns about unequal access to advanced AI tools were also commonly reported, with 44.73% (n=297/664) of respondents agreeing. Notably, 38.01% (n=252/663) of respondents strongly agreed that GenAI chatbots raise data privacy and confidentiality issues.

**Figure 3.**
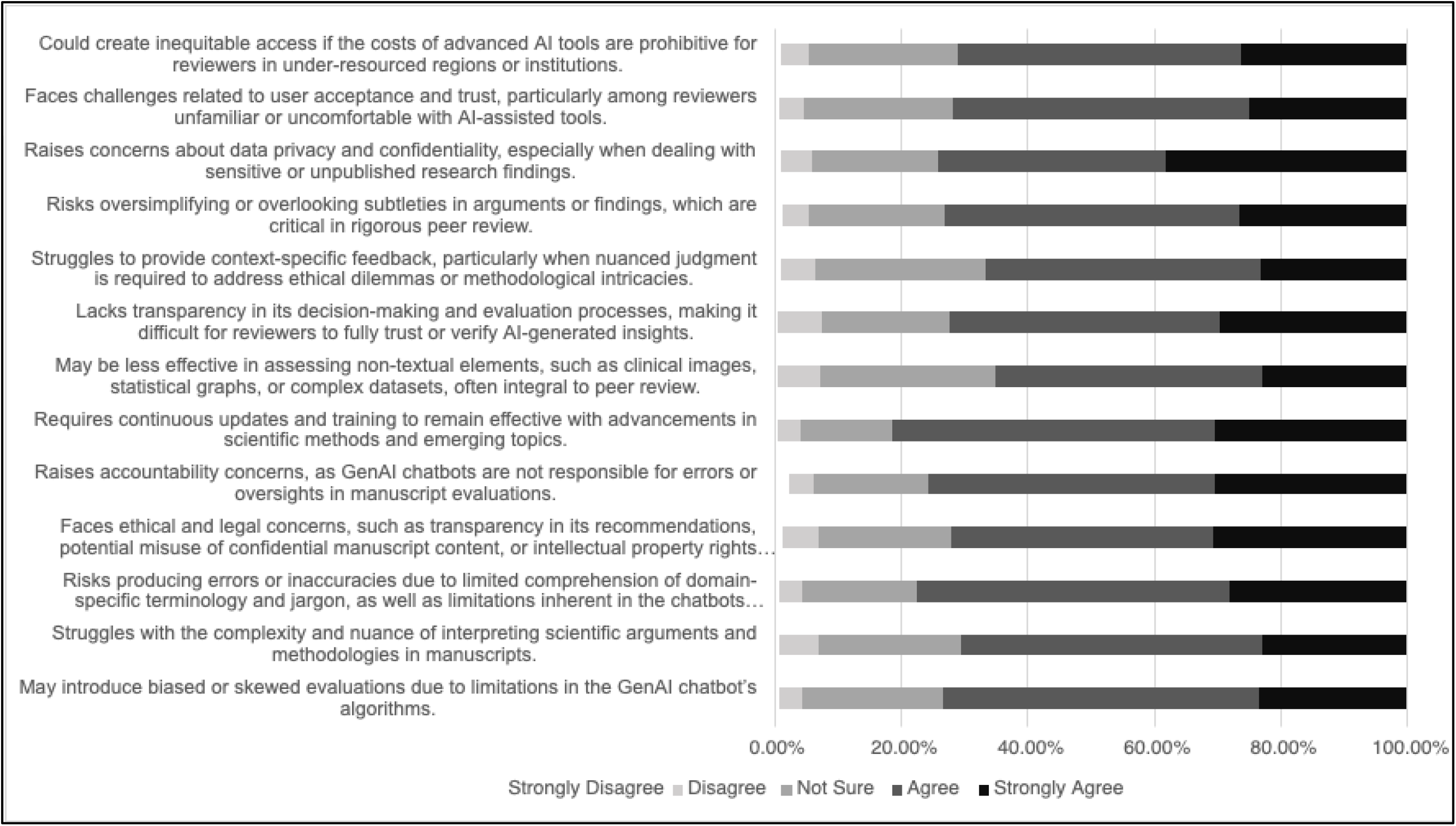
TCIM Peer Reviewers’ Perceptions of the Proposed Challenges of GenAI Chatbot Use in the Peer Review Process.

Thematic analysis of the open-ended responses indicated a general understanding among respondents that GenAI chatbots can be useful tools to assist the peer review process, provided they are employed within well-defined ethical, professional, and institutional boundaries. Across responses, participants consistently emphasized that GenAI should function as a supplementary aid rather than a replacement for the expertise, judgment, and accountability of human peer reviewers. Three overarching themes emerged: perceived benefits of GenAI, concerns regarding accuracy and research integrity, and the need for governance, training, and human oversight. The complete thematic coding framework and supporting data are available in the supplementary files on OSF at https://osf.io/7vqj4/files/nzc7q.

## Discussion

The primary objective of this study was to examine TCIM peer reviewers’ attitudes and perceptions toward the use of GenAI chatbots in the peer-review process. Overall, most respondents expressed cautiously positive views regarding the future role of GenAI chatbots in peer review, while simultaneously acknowledging important ethical, methodological, and practical concerns associated with their use.

## Comparative Literature

To our knowledge, no previous studies have specifically examined perceptions of GenAI among TCIM peer reviewers. Nevertheless, our findings are largely consistent with those of our companion study exploring TCIM researchers’ perceptions of GenAI across the broader scientific process, as well as with the emerging literature on AI-assisted scholarly publishing [30]. Most respondents in the present study were familiar with GenAI chatbots, with OpenAI’s ChatGPT being the most widely used platform, and generally viewed these technologies as most useful for supporting language-related and administrative tasks, including summarizing manuscripts, improving grammar and readability, organizing reviewer notes, and assisting with drafting constructive feedback. Participants frequently agreed that the primary benefits of GenAI-assisted peer review were increased efficiency and faster completion of peer reviews (50.72%), reduced reviewer burden (51.16%). They improved the clarity and readability of peer review reports (48.33%). These findings are consistent with previous studies suggesting that large language models may improve the efficiency and quality of scholarly publishing activities. Similarly, Liang et al. demonstrated that large language models can provide useful feedback on research manuscripts, particularly regarding writing quality and reporting [34].

Despite recognizing these potential benefits, respondents also expressed considerable concerns regarding the use of GenAI during peer reviews. The most commonly reported concerns were algorithmic bias (49.85%) and AI systems’ inability to adequately interpret complex scientific reasoning and research methodologies (47.67%). A large proportion of participants agreed that GenAI raises ethical and legal concerns related to transparency, confidentiality, and consent (41.38%). Qualitative responses emphasized concerns about the confidentiality risks of uploading unpublished manuscripts to external AI systems. These findings are consistent with broader concerns in the literature about the limitations of large language models in contexts that require disciplinary expertise, contextual interpretation, and independent scientific decision-making. Likewise, Hosseini and Horbach argued that although GenAI may help alleviate reviewer burden, its use may introduce new risks to research integrity if implemented without appropriate safeguards [1].

Notably, this study identified a discrepancy between respondents’ perceptions of the future importance of GenAI and their willingness to adopt these technologies in peer review. Although a vast majority of respondents believed GenAI would become increasingly relevant within the peer review process, only approximately half indicated that they intended to use these tools in their future reviewing practices. This discrepancy may reflect concerns about appropriate implementation, limited trust in existing GenAI systems, and a lack of clear institutional guidance on their use. Consistent with this interpretation, relatively few respondents reported that their institutions had established policies regarding GenAI-assisted peer review. At the same time, most agreed that reviewers would benefit from education and training to facilitate responsible use of these technologies. Given that GenAI represents a relatively recent development in scholarly publishing, these findings likely reflect the current lack of standardized guidance for its responsible use.

These concerns are also reflected within the broader scholarly publishing community. Although many journals, publishers, and professional organizations have begun implementing policies governing the use of GenAI in peer review, issues remain regarding undisclosed AI use, misinformation, data leakage, accountability, and fabricated citations [35, 36]. Consistent with these concerns, publishers, journals, and organizations have introduced guidance emphasizing confidentiality, transparency, disclosure of AI use, and the continued accountability of human reviewers during AI-assisted peer review [37, 38]. Collectively, these findings suggest that the successful integration of GenAI into TCIM peer review will depend not only on technological capability but also on the development of clear institutional policies, reviewer education, and robust ethical frameworks. These challenges are already emerging in editorial practice, as unusually rapid turnaround and prolific reviewing activity can raise suspicions of undisclosed AI use and questions about how potential misuse should be addressed and detected [37, 39].

Finally, TCIM peer reviewers may hold unique perspectives that differ from those of reviewers in other scientific disciplines because of the multidisciplinary, culturally grounded, and heterogeneous nature of TCIM evidence systems and methodologies. Peer review within TCIM often involves evaluating complex interventions, traditional knowledge systems, and context-dependent therapeutic approaches that may not be adequately represented within current GenAI training data. Consequently, respondents may be particularly cautious regarding the ability of GenAI chatbots to appropriately interpret nuanced scientific reasoning, cultural context, and methodological diversity within TCIM scholarship. At the same time, participants recognized that GenAI may offer opportunities to reduce reviewer fatigue, improve interdisciplinary communication, and support the growing demands placed on peer review processes. Collectively, with findings from our companion study [30], these results suggest that TCIM researchers generally view GenAI as a valuable supportive tool that should complement, rather than replace, human expertise throughout the peer review process [30].

### Strengths and Limitations

This study had several notable strengths, including its large international sample and inclusion of both quantitative and qualitative data exploring perceptions of GenAI chatbot use in peer review. Conducting a cross-sectional survey also enabled data collection at a single point, making the study cost-effective. By employing a sampling strategy that sought a large, geographically diverse international participant pool, we captured perspectives from TCIM reviewers across multiple countries and research settings, thereby increasing the scope and potential generalizability of our findings.

However, there are some limitations. The high sampling frame was accompanied by a relatively low response rate, which could indicate nonresponse bias. This may have been attributed to outdated contact information, inaccessible email addresses, changes in contact affiliations, bounce-back emails, vacations, retirement from work, and unfamiliarity with the researchers conducting the study or their lack of anticipation of receiving an invitation to participate in the survey. Additionally, individuals with greater interest in or familiarity with GenAI technologies may have been more likely to participate, potentially resulting in responses that are more informed or favorable than those of the broader TCIM peer reviewer population.

Furthermore, although not all corresponding authors of TCIM research are necessarily involved in peer review, a considerable overlap between researchers and peer reviewers can reasonably be assumed, given that active researchers are commonly recruited as peer reviewers based on their publication expertise. The survey assessed institutional policies but did not examine the journal or publisher policies governing respondents’ peer-review activities. Consequently, we could not determine if the inviting journal or publisher permitted GenAI, whether reviewers sought permission, or if such use was disclosed. This limits our ability to interpret reported practices in the context of editorial policies. Future studies should distinguish between institutional and journal and publisher governance and assess reviewers’ awareness of and disclosure under these policies. Although respondents were free to report experiences with any GenAI chatbot system, participation may have been limited in certain regions because SurveyMonkey is inaccessible in some countries and because peer reviewers in some regions may have had limited English proficiency, potentially reducing representation from these academic communities. Future studies may benefit from offering multilingual surveys or alternative survey platforms to improve accessibility and increase participation from underrepresented regions. Moreover, recall bias may have influenced the results, as participants might not have recalled their prior use of GenAI chatbots during peer reviews. Finally, because of the cross-sectional design of this study, the findings represent perceptions at a single point in time. Given the rapid development of GenAI technologies, institutional policies, and publisher guidance, attitudes toward GenAI-assisted peer review may continue to evolve.

## Conclusion

Overall, this study provides the first large-scale international examination of TCIM peer reviewers’ perceptions of the use of GenAI chatbots in peer review. Although respondents recognized the potential of GenAI to improve efficiency, reduce reviewer burden, and support language-related tasks, these perceived benefits were tempered by concerns regarding bias, transparency, and confidentiality. Importantly, while most respondents viewed GenAI as important to the future of peer review, only about half reported being likely to use these tools, highlighting a gap between perceived importance and willingness to adopt. These findings are particularly relevant to TCIM, where peer review often involves evaluating culturally grounded knowledge systems and diverse methodological approaches. Respondents appeared more accepting of GenAI for supportive and administrative tasks than for evaluating scientific rigour or research integrity, reflecting concerns that current GenAI systems may not adequately capture the contextual and epistemological nuances specific to TCIM research. Collectively, these findings suggest that the responsible integration of GenAI into TCIM peer review will require clear policies, reviewer education, and robust ethical safeguards. Most importantly, GenAI should serve as a supportive tool that complements human expertise and judgment in the peer-review process.

## Data Availability

All relevant study materials and data are included in this manuscript or posted on the Open Science Framework.

https://doi.org/10.17605/OSF.IO/M6SGP

## List of Abbreviations

AI: artificial intelligence
CHERRIES: checklist for reporting results of internet e-surveys
DOI: digital object identifier
GenAI: generative artificial intelligence
NCCIH: National Center for Complementary and Integrative Health
OSF: Open Science Framework
PMID: PubMed Identifier
STROBE: Strengthening the Reporting of Observational Studies in Epidemiology
TCIM: traditional, complementary, and integrative medicine
WHO: World Health Organization

## Declarations

### Ethics Approval and Consent to Participate

Ethics approval was sought and obtained from the University Hospital Tübingen Research Ethics Board (REB #: 543/2025BO2) to conduct this study.

### Consent for Publication

All authors consent to this manuscript’s publication.

### Availability of Study Materials and Data

All relevant study materials and data are included in this manuscript or posted on the Open Science Framework: https://doi.org/10.17605/OSF.IO/M6SGP

### Authors’ Contributions

JYN: designed and conceptualized the study, collected and analysed data, drafted the manuscript, and gave final approval of the version to be published.

JT: assisted with the collection and analysis of data, made critical revisions to the manuscript, and gave final approval of the version to be published.

KA: assisted with the collection and analysis of data, made critical revisions to the manuscript, and gave final approval of the version to be published.

PKG: assisted with the collection and analysis of data, made critical revisions to the manuscript, and gave final approval of the version to be published.

MG: assisted with the collection and analysis of data, made critical revisions to the manuscript, and gave final approval of the version to be published.

SL: assisted with the collection and analysis of data, made critical revisions to the manuscript, and gave final approval of the version to be published.

DM: assisted with the collection and analysis of data, made critical revisions to the manuscript, and gave final approval of the version to be published.

MR: assisted with the collection and analysis of data, made critical revisions to the manuscript, and gave final approval of the version to be published.

MS: assisted with the collection and analysis of data, made critical revisions to the manuscript, and gave final approval of the version to be published.

JPS: assisted with the collection and analysis of data, made critical revisions to the manuscript, and gave final approval of the version to be published.

TY: assisted with the collection and analysis of data, made critical revisions to the manuscript, and gave final approval of the version to be published.

MSL: assisted with the collection and analysis of data, made critical revisions to the manuscript, and gave final approval of the version to be published.

HC: assisted with the collection and analysis of data, made critical revisions to the manuscript, and gave final approval of the version to be published.

